# Protective effect of BNT162b2 vaccination on aerobic capacity following mild to moderate SARS-CoV-2 infection: a cross sectional study, Israel, March-December 2021

**DOI:** 10.1101/2021.12.30.21268538

**Authors:** Yair Blumberg, Michael Edelstein, Kamal Abu Jabal, Ron Golan, Yuval Perets, Musa Saad, Tatyana Levinas, Saleem Dabbah, Zeev Israeli, Salah Yacoubi, Alaa Abu Raya, Anat Amital, Majdi Halabi

## Abstract

There is increasing evidence that patients who were infected with SARS-CoV-2 may experience adverse health outcomes months after the acute infection has resolved including reduction in aerobic capacity and fatigue. In this study, we compared aerobic capacity and exercise performance of 28 unvaccinated participants to 15 vaccinated ones who performed a symptom limited cardio-pulmonary exercise test (CPET) after acute COVID-19. We identified a significant difference in aerobic capacity between vaccinated and unvaccinated individuals, with a lower V’O2 peak percentage of predicted in the unvaccinated group. In addition, the unvaccinated group had a reduction in the peak-exercise heart rate and lower ventilation values. Our results suggest objective limitations to exercise capacity in the months following acute COVID19 illness, mitigated by vaccination

## Introduction

Patients who were infected with acute respiratory syndrome coronavirus 2 (SARS-CoV-2) may experience post-acute adverse health outcomes, a phenomenon that has been named long COVID. The current case definition remains very broad [1] but the most commonly reported symptoms are fatigue, headache and attention/concentration issues, with dyspnoea being fifth [2]. In addition, reduced aerobic capacity has been demonstrated in both mild and moderate COVID19 patients[3].

The impact of vaccination on long COVID remains unclear. One large British study showed an association between COVID19 vaccination and a reduction in reported post-acute symptoms, without specifying the symptoms or follow-up duration [4]. Another study showed that one or more doses of COVID-19 vaccine was not associated with a reduction in reported long-COVID symptoms, but two doses were [5]. These studies measure self-reported symptoms. This study is, to our knowledge, the first to compare vaccinated and unvaccinated individuals previously infected with SARS-CoV-2 in terms of aerobic capacity and exercise performance.

## Methods

This prospective cross-sectional study was conducted at the cardiac rehabilitation department of Ziv Medical Centre, a 300-bed government hospital in Safed, Northern Israel, between March 2021 and December 2021 (Ethics approval:0100-20-ZIV). Individuals aged 18 to 65 years with previously documented mild to moderate COVID-19 disease were eligible to participate. Each participant performed a symptom-limited cardio-pulmonary exercise test (CPET) using an individually calibrated bicycle ergometer (Cortex-Medical). Prior to exercise, each patient underwent spirometry tests according to the American Thoracic Society protocol [6]. The CPET test was performed on a cycle ergometer; subjects were asked to maintain a constant pedalling frequency of 60 ± 5 revolutions per minute. Throughout the test, cardiac electrical activity was monitored using continuous electrocardiography. Blood pressure and Perceived Exertion (RPE) were measured every two minutes.

Peak oxygen consumption (V’O_2_) was defined as the highest value of V’O_2_ attained in a 20 second interval. Age-predicted heart rate (HR) was calculated as 220-age (bpm). The anaerobic threshold (AT), referring to the point at which ventilation starts to increase at a faster rate than V’O_2_, was determined by the V-slope method [6].

The minute ventilation/carbon dioxide production (V’E/V’CO_2_) slope was calculated as the coefficient of linear regression obtained by plotting the V’E and V’CO_2_ data of the subject’s exercise phase.

We described the characteristics of each group and compared them using chi square tests. We calculated means and standard errors (SE) for each observation category. We compared observed CPET results to predicted values within each group using t-tests. Between-group differences were measured using chi square (for proportions) and t-tests (for means). Analysis was performed using SPSS software (IBM SPSS Statistics for Windows, Version 25.0. Armonk, NY). P-value of 5% or less was considered statistically significant.

## Results

Forty-three participants were enrolled: 28 unvaccinated and 15 vaccinated; patient characteristics and symptoms during and after the acute COVID-19 episode are presented in Table 1. All patients were infected prior to the Omicron variant being imported to Israel. All vaccinated patients except for 2 received at least 2 doses, and all were infected post vaccination. The CPET test was conducted at a mean of 119±24 days after acute disease with no difference between the two groups (p=0.12). There was no difference in baseline demographic and pulmonary function test (PFT) parameters, six participants from the unvaccinated group had comorbidities as presented in Table 1. The major cardiopulmonary metrics are summarized in Table 2. Compared with unvaccinated individuals, those vaccinated had higher mean V’O_2_/kg at peak exercise as well as mean peak heart rate (HR) (Figure 1). The mean V’O_2_/kg peak in the unvaccinated group was 83% of predicted vs 95% in the vaccinated (p<0.05, Table 2); In the unvaccinated group, 14/28 subjects (50%) had a V’O_2_ peak < 80% of predicted vs 2/15 (14%) among those vaccinated. The maximum HR and VE were reduced among unvaccinated participants compared with those vaccinated (Figure-1, Table 2).

**Table 1.** Participant characteristics, symptoms and pulmonary function test (PFT)

|  | Unvaccinated (n=28) | Vaccinated (n=15) |
| --- | --- | --- |
| Male (n, %) | 15 (53%) | 7 (46%) |
| Female (n, %) | 13 (46%) | 8 (53 %) |
| Age (years, SE) | 46.6±12 | 9±39.8 |
| Height, (cm, SE) | 170±8 | 13±169 |
| Weight,( kg, SE) | 82.5±16 | 12±77.5 |
| Body Surface Area | 1.8±0.6 | 0.22±1.92 |
| Body mass Index | 28±6 | 3.2±26.8 |
| Comorbidities |  |  |
| Diabetes Mellitus (n, %) | 4 (14%) | - |
| Hypertension (n, %) | 4 (14%) | - |
| PFT (Pulmonary Function Test) |  |  |
| Forced Vital Capacity (FVC) | 4.5±1.2 | 1.5±4.8 |
| Forced Expiratory Volume (FEV<sub>1</sub>) | 3.5±0.9 | 1.1±3.8 |
| FEV<sub>1</sub>/FVC | 78.5±8 | 7.2±79.5 |
| Symptoms during acute COVID-19 |  |  |
| Dyspnea | 15 (53%) | 4 (26%) |
| Cough | 10 (35%) | 6 (40%) |
| Runny nose | 3 (10%) | 4 (26%) |
| Loss of taste or smell | 13 (46%) | 10 (66%) |
| Headache | 16 (57%) | 8 (15%) |
| Dizziness | 4 (14%) | 1 (6%) |
| Diarrhea | 1 (3%) | 1 (6%) |
| Muscle or body aches | 19 (67%) | 11 (73%) |
| Fever | 14 (40%) | 5 (33%) |
| Post-acute symptoms |  |  |
| Fatigue | 14 (40%) | 5 (33%) |
| Muscle or body aches | 8 (28%) | 2 (13%) |
| Effort dyspnea | 15 (53%) | 5 (33%) |
| loss of taste or smell | 5 (17%) | 2 (13%) |

**Table 2.**
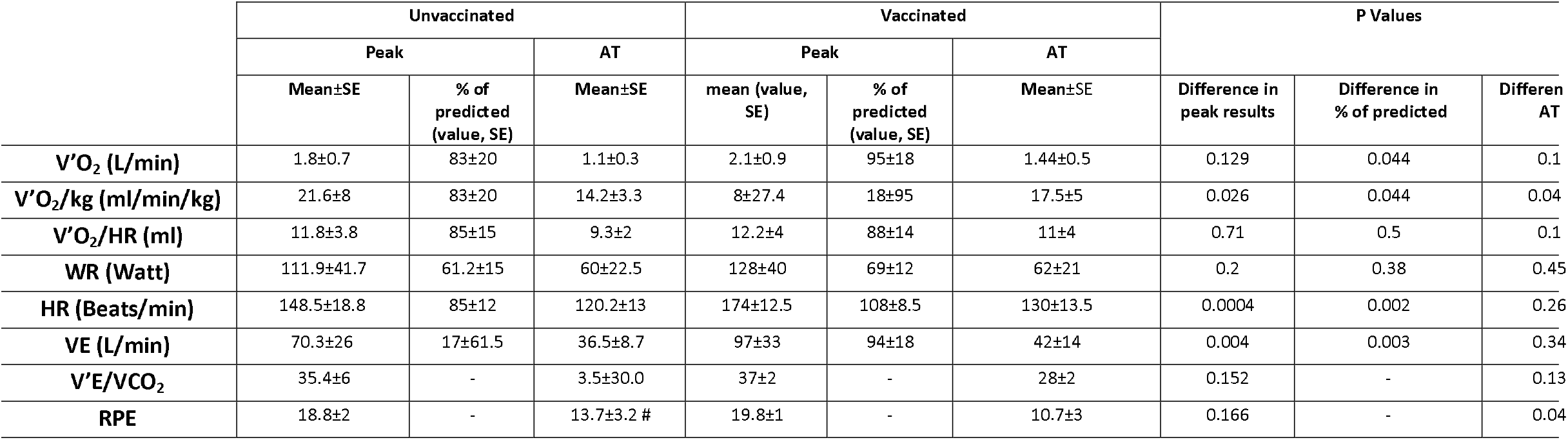
CPET parameters at peak exercise and AT among vaccinated and unvaccinated patients previously infected with COVID19

**Figure 1:**
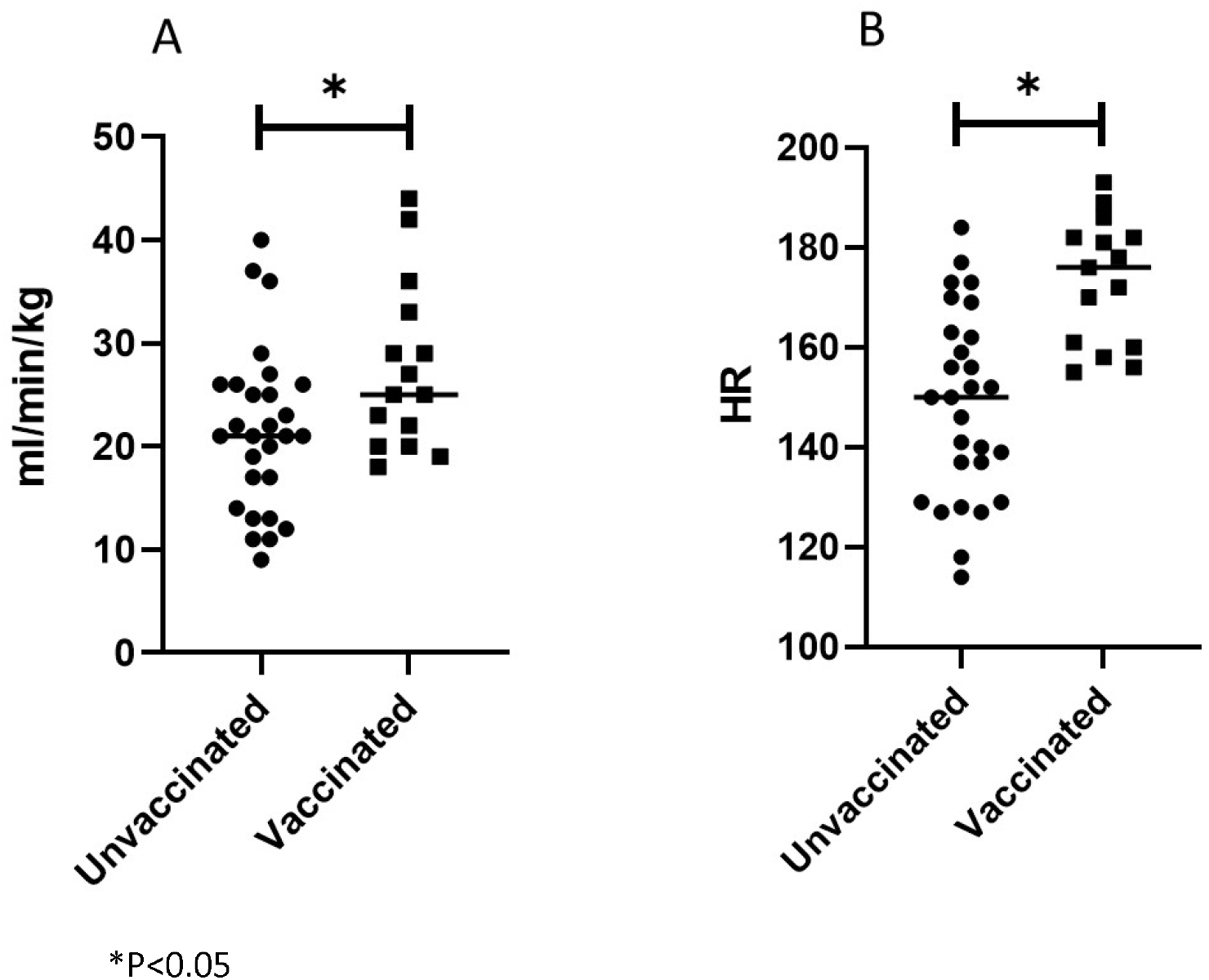
VO_2_/Kg peak (panel A) and Peak Heart Rate (panel B) among COVID19 Unvaccinated vs Vaccinated patients previously infected with SARS-CoV-2

## Discussion

This study aimed to determine whether vaccination was associated with exercise capacity as measured with CPET among individuals previously infected with SARS-CoV-2, after the acute phase of COVID-19.

We found a difference in aerobic capacity between vaccinated and unvaccinated individuals previously infected with SARS-CoV-2. Unvaccinated individuals on average reached a lower proportion of their predicted peak V’O_2,_ and HR. A high proportion of unvaccinated individuals performed poorly (<80% of predicted) on these indicators.

These results suggest chronotropic incompetence in the unvaccinated group, with a reduction in the peak-exercise HR and VE, contributing to limited exercise capacity. Similar results have been demonstrated for HR in other studies [7,8]. In our cohort we show that there is also a difference in ventilation (V’E), with lower values for the unvaccinated group.

Identifying the pathological mechanism leading to an inability to increase HR and ventilation are beyond the scope of this study. Suggested mechanisms in the literature include Immune-mediated damage to the autonomic nervous system during COVID-19 and a peripheral cardiac limit to exercise resulting from an oxygen diffusion defect [9,10]

### Limitations

Our study has some limitations, including the small cohort size and differences in baseline characteristics. However, the fact that we measure findings in individuals against their own predicted value adjusts for these differences to a large extent. A sub analysis excluding patients with co-morbidities did not change the findings (data not shown). Two of the patients were only partially vaccinated, possibly leading to a slight underestimation of the effect of vaccination. We did not evaluate blood gas, which could have revealed more about the main cause of exercise limitation for patients with reduced pVO_2_.

## Conclusion

This study suggests that COVID19 patients can suffer from objective limitations to exercise capacity in the months following their acute episode. Our study is the first to show a protective effect of vaccination against decreased aerobic capacity. As a more objectively quantifiable definition of long COVID is needed, studies able to demonstrate measurable changes post-acute infection with SARS-CoV-2 are essential. The measured protective effect of vaccination gives additional reasons to continue and intensify the vaccine drive globally and similar studies should be replicated on a larger scale to confirm our results.

## Data Availability

All data produced in the present work are contained in the manuscript

## Funding statement

the study received no external funding

## Conflict of interest statement

the authors declare no conflict of interest

## Notes

### Competing Interest Statement

The authors have declared no competing interest.

### Funding Statement

This study did not receive any external funding

### Author Declarations

The ethics committee of Ziv Medical Centre approved the study, approval 0100-20-ZIV

### Summary of Updates

Correction of a number in the abstract and use of the corrected abstract version

